# Plasma and neuroimaging biomarkers of small vessel disease and Alzheimer’s disease in a diverse cohort: MESA

**DOI:** 10.1101/2025.02.11.25322109

**Authors:** Samuel N. Lockhart, Courtney L. Sutphen, Jordan Tanley, Fernando Gonzalez-Ortiz, Przemysław R. Kac, Mohamad Habes, Susan R. Heckbert, Nicholas J. Ashton, Michelle M. Mielke, Robert Koeppe, Marc D. Rudolph, Christopher T. Whitlow, Kevin D. Hiatt, Suzanne Craft, Thomas C. Register, Kathleen M. Hayden, Stephen R. Rapp, Bonnie C. Sachs, Henrik Zetterberg, Kaj Blennow, Thomas K. Karikari, Timothy M. Hughes

## Abstract

**INTRODUCTION:** Little is known about how Alzheimer’s disease (AD) plasma biomarkers relate to cerebral small vessel disease (cSVD) neuroimaging biomarkers.

**METHODS:** The study involved 251 Wake Forest Multi-Ethnic Study of Atherosclerosis (MESA) Exam 6 participants with plasma AD biomarkers, MRI, amyloid PET, and adjudicated cognitive status. Multivariable models examined cross-sectional relationships between plasma and neuroimaging biomarkers, considering comorbidities.

**RESULTS:** Lower Aβ42/Aβ40, and higher GFAP, NfL, and p-tau217 were associated with greater neurodegeneration. Lower plasma Aβ42/Aβ40 and higher p-tau217 and p-tau231 were associated with greater Aβ PET deposition. NfL was positively associated with WMH and WM Free Water. P-tau measures were positively associated with WM Free Water. Lower Aβ42/Aβ40 was associated with presence of microbleeds. GFAP was positively associated with WMH.

**DISCUSSION:** We observed expected associations of plasma biomarkers with cognitive status and imaging biomarkers. GFAP, NfL, p-tau181, p-tau217, and p-tau231 are associated with cSVD in addition to AD-related pathology.

## Introduction

Plasma biomarkers have great potential to inform disease etiology in Alzheimer’s disease (AD) and related dementias (ADRD). Selected plasma biomarkers of amyloid, phosphorylated tau (p-tau), and neurodegeneration detect AD pathophysiology, monitor disease course, and predict future risk for ADRD—some more accurately than others^1–6^. Consequently, these biomarkers are being included in research and clinical programs^7,8^, often alongside existing MRI and PET measures of AD/ADRD pathological changes^9–11^. Several studies report associations of plasma AD biomarkers with vascular, metabolic and renal comorbidities^12,13^. In addition, plasma p-tau levels increase with acute neuronal injury, such as cardiac arrest^14^ and traumatic brain injury^15^, which may be related to release across the blood-brain barrier in the cerebral microvessels. However, few studies assessing plasma AD biomarkers have evaluated measures of cerebral small vessel disease (cSVD, e.g., brain microbleeds and white matter injury) common in older adults^16–20^. Biomarkers of cSVD are key for understanding the vascular contributions to cognitive impairment and dementia in the context of AD/ADRD^21^.

Importantly, blood and imaging biomarkers of AD/ADRD have been mostly studied in non-Hispanic White (NHW) individuals, with limited data on diversity in race, ethnicity, and socioeconomic factors^8^. Some recent reports suggest racial and ethnic differences in the ability of blood AD biomarkers to predict abnormal brain amyloid load^22^, consistent with observed racial and ethnic differences in cerebrospinal fluid biomarkers^23–31^. Yet, most of these studies do not investigate the consistency of relationships between plasma AD biomarker levels and other biomarkers of AD and cSVD among various racial and ethnic groups. This is critically important because underrepresented groups tend to have a higher prevalence of cardiometabolic comorbidities contributing to cerebral small vessel disease (e.g., diabetes, hypertension, heart, and kidney diseases) than NHW participants.

Further, comorbidities can affect blood AD protein biomarker production and clearance mechanisms^32,33^. Recently, Mielke et al.^34^ reported differences in plasma p-tau biomarkers by these comorbidities in a NHW cohort. It is important to understand how comorbidities affect plasma AD biomarkers in other populations with different rates of age-related comorbidities^13^. Indeed, the higher prevalence of these comorbidities among racial and ethnic populations may contribute to group differences in AD biomarker levels and result in inaccurate diagnoses^34,35^.

The Multi-Ethnic Study of Atherosclerosis (MESA) is a unique, diverse study with over 20 years of extensive longitudinal vascular phenotyping as well as cognitive testing^36,37^. During MESA Exam 6, a diverse (White and Black/African-American) cohort at the Wake Forest University (WFU) site underwent brain MRI, amyloid PET, and assessment of plasma AD biomarkers. Here, we leveraged data from the WFU site of MESA Exam 6 to examine cross-sectional relationships between plasma AD biomarkers (amyloid-beta [Aβ42/Aβ40 ratio], glial fibrillary acidic protein [GFAP], neurofilament light chain [NfL], p-tau181, p-tau217, and p-tau231), comorbidities, and neuroimaging biomarkers of cSVD and AD/ADRD among individuals self-reporting as NHW and Black/African American. We hypothesized that we would observe associations of plasma AD biomarkers with imaging measures of brain amyloid PET, cSVD, and neurodegeneration.

## Methods

### Participants

MESA participants were recruited from six field centers and free from clinical cardiovascular disease (including stroke) at baseline (2000-02)^36,37^. Only participants at the WFU field center at Exam 6 (2016-18) were included in this analysis. Demographic, anthropometric, and standard clinical data were collected in MESA as previously reported, including baseline years of education, self-reported gender, and self-reported race and ethnicity (viewed here as a social construct), as well as Exam 6 age, smoking status, and height and weight for body mass index (BMI). Estimated glomerular filtration rate (eGFR, in mL/min/1.73m^2^; Exam 6) was calculated using the creatinine-based four-variable Modification of Diet in Renal Disease (MDRD) equation^38^ with no race adjustment. DNA from baseline was analyzed for *APOE* genotypes as previously described^40^; *APOE*-ε4 carriage was defined as presence of one or more ε4 allele(s). The research protocol was approved by the local Institutional Review Board, with informed consent obtained for all participants.

### Cognitive testing and adjudication

MESA Exam 6 participants at WFU field center were administered the Uniform Data Set version 3 (UDSv3) neuropsychological battery^41^, including: detailed cognitive testing; ratings of functional abilities by a person familiar with the participant; information about family history of AD, medications, and health history; clinician-assessed medical conditions and judgment of symptoms; and a neurological examination. All of these data were used in the adjudication process along with appropriate normative data^38^. The consensus panel consisted of neuropsychologists, geriatricians, neurologists, and other aging experts. Consensus-adjudicated cognitive status, according to published criteria^39,40^ included cognitively unimpaired (CU), mild cognitive impairment (MCI), and dementia.

### Plasma biomarker acquisition and processing

Plasma AD biomarker analysis was conducted from stored plasma samples from WFU field center participants at Exam 6. Plasma AD biomarker assay results of amyloid-beta (Aβ42, Aβ40, and their ratio), glial fibrillary acidic protein (GFAP), neurofilament light chain (NfL), p-tau181, p-tau217, and p-tau231. All biomarkers were measured on the Quanterix Single molecule array (Simoa) HD-X platform (Quanterix, Billerica, MA) with a twofold dilution factor in plasma.

Plasma Aβ42, Aβ40, NfL, and GFAP concentrations were measured using commercially available Neurology 4-Plex E kits on the HD-X. Plasma p-tau181^2^, p-tau231^4^, and University of Gothenburg (UGOT) p-tau217^43^ concentrations were measured on HD-X with methods published previously. Signal variations within and between analytical runs were assessed using three internal quality control samples at the beginning and the end of each run.

### MRI acquisition and processing

Brain MRI data were acquired for participants at WFU on a 3T Siemens Skyra scanner using a high-resolution 32-channel head coil. T1 MPRAGE (to quantify gray matter [GM] volume and cortical thickness), and T2 fluid-attenuated inversion recovery (FLAIR; to quantify white matter [WM] hyperintensities [WMH]) were acquired; sequence details have been published previously^44^. Neurite orientation density and dispersion imaging (NODDI; 2.0 mm isotropic resolution, TR=3500 ms, TE=106 ms, FA=90, b=714/1000/2855 s/mm^2^, 131 directions) was acquired to examine isotropic volume fraction (i.e., “free water” [FW]). Susceptibility Weighted Imaging/Quantitative Susceptibility Mapping (SWI/QSM) images were acquired (TR=51 ms; multiple TE=9.8, 16.6, 23.5, 30.4, 37.3, 44.2 ms; FA=20; 0.6 x 0.6 x 2mm) to enable visualization of microbleeds.

MRI data for this analysis were processed at the WFU field site. Regional volumes and cortical thicknesses were calculated on T1 using FreeSurfer v5.3 (https://surfer.nmr.mgh.harvard.edu). We examined neurodegeneration using bilateral hippocampal volume (HCV) divided by FreeSurfer total intracranial volume (ICV; to correct for head size), as well as total GM volume (GMV) divided by ICV. We additionally calculated cortical thickness in a temporal lobe region of interest (ROI) shown to index neurodegeneration in regions characteristic of age-related dementias; this was calculated by averaging surface area-weighted cortical thickness of bilateral entorhinal, inferior/middle temporal, and fusiform regions^45^. WMH lesions were segmented as described previously^44^ by the lesion growth algorithm (LGA) implemented in LST toolbox v2.0.15 (www.statistical-modelling.de/lst.html), running in MATLAB SPM12 (www.fil.ion.ucl.ac.uk/spm) using FLAIR with T1 as reference.

Total WMH lesion volume was divided by FreeSurfer ICV and log-normalized to generate a global WMH volume measure (lnWMH). NODDI processing details have been described previously^46^; briefly, the Johns Hopkins University (JHU) DTI atlas was overlaid on template-space FW images to extract mean signal across all supratentorial WM tracts to calculate mean global WM FW, and a set of all supratentorial Automated Anatomical Labeling (AAL) gray matter (GM) ROIs were overlaid on template-space FW images to calculate mean global GM FW. A single trained neuroradiologist (KDH) read cerebral microbleeds and lacunar infarctions according to STRIVE criteria^47^ (on n=236 participants with available imaging), which were binarized into presence/absence. For purposes of these analyses, we classified MRI-based biomarkers into measures of neurodegeneration/neuroinflammation (HCV, GMV, Cortical Thickness, GM FW) and cSVD (lnWMH, WM FW, cerebral microbleeds, lacunar infarction).

### PET acquisition and processing

On a subset of n=177 participants, [^11^C] Pittsburgh Compound B (PiB)^48^ was used for assessing fibrillar amyloid brain deposition on PET, using acquisition methods described previously^44^.

Following a computed tomography (CT) scan for attenuation correction, participants were injected with ∼370 MBq [^11^C]PiB and scanned 40–70 min (6 × 5-min frames) post-injection on a 64-slice GE Discovery MI DR PET/CT scanner.

Centiloid (CL)-based processing was conducted by the University of Michigan to generate global CL values (whole cerebellum reference region, 50-70 min data)^49^. We examined continuous global CL values; additionally, a CL value of 12.2, which represents CERAD moderate-to-frequent neuritic plaques^50^, was used as a primary threshold for Aβ PET positivity.

### Statistical Analysis

This analysis was limited to 251 MESA Exam 6 participants at the WFU field center with MRI data, plasma AD biomarkers, and UDSv3-based adjudicated cognitive status. Chi square and ANOVA tests were used to examine differences in baseline demographics by cognitive classification. Prior to analysis in regression models, we standardized the distribution of each plasma AD biomarker by log2 transformation, where parameter estimates represent a doubling of each biomarker level.

We used multivariable general linear models (GLM) to assess relationships between plasma AD biomarkers (Aβ42/Aβ40 ratio, GFAP, NfL, p-tau181, p-tau217, p-tau231) and MRI measures of brain small vessel disease and neurodegeneration (GMV, HCV, Cortical Thickness, lnWMH, GM FW, WM FW), and PET measures of global amyloid deposition (Aβ-positivity; 12.2 CL value). Prior to construction of GLMs, we assessed collinearity among neuroimaging-based outcome variables. MRI brain volume estimates were corrected for total intracranial volume; global WMH volume was also log-transformed prior to analysis. For amyloid PET, we also examined the relationship with a 24.4 CL threshold, representing intermediate to high AD neuropathological changes^50^ and found identical results (not shown) for AD plasma biomarkers as we did for the 12.2 CL threshold.

Model 1 included basic covariates of age, gender, race, and education. Model 2 included covariates from model 1, plus *APOE*-ε4 carrier status, smoking status, and comorbidities associated with plasma AD biomarker levels (BMI, eGFR; both treated as continuous variables when used as model covariates). Each analysis examined effect modification by comorbidities (BMI, eGFR), age (median split), race, gender, and *APOE*-ε4 status. We also conducted a sensitivity analysis excluding n=44 participants with severe chronic kidney disease (CKD), defined using eGFR≤60 mL/min/1.73m^2^).

We examined consistency of relationships of plasma AD biomarkers with MRI measures of brain small vessel disease and neurodegeneration and amyloid positivity on PET among self-reported White and Black older adults after considering differences in comorbidities. We further explored the impact of medical comorbidities, such as BMI and kidney function (assessed with estimated glomerular filtration rate, eGFR), on plasma AD biomarker levels^13^. Models were corrected for multiple comparisons using the Benjamini-Hochberg False-discovery rate (FDR)^51^.

## Results

### Participants

Demographics by cognitive status at Exam 6 for the 251 participants in the present study are presented in **Table 1**, with 69% adjudicated as CU, 27% with MCI, and 4% with probable dementia. Cognitive status groups differed by age, gender, and race. They also differed by eGFR and *APOE*-ε4 carrier status, such that (continuous) eGFR was lower and the prevalence of *APOE*-ε4 carrier status was higher in MCI and dementia groups. Those with MCI or dementia demonstrated a greater degree of neurodegeneration, with lower GMV, lower HCV, lower cortical thickness, and higher GM FW. In the subset of participants with amyloid PET data, amyloid positivity was higher in MCI and dementia groups, compared to CU participants. There were no differences across cognitive groups in measures of vascular injury (WMH volume, WM Free Water, presence or number of microbleeds, and presence of lacunes). **Supplementary Table 1** describes associations between plasma AD biomarker levels and covariates included in models.

**Table 1.** Demographic and Clinical Characteristics by Cognitive Status

|  |  | CU |  | MCI |  | Dementia |  |  |
| --- | --- | --- | --- | --- | --- | --- | --- | --- |
|  |  | (n = 172) |  | (n = 69) |  | (n = 10) |  |  |
|  |  | N / Mean | % / SD | N / Mean | % / SD | N / Mean | % / SD | P-value |
| Age (y) |  | 71.6 | 6.9 | 74.4* | 7.2 | 78.6* | 4.9 | <0.001 |
| Education (y) |  | 15.3 | 2.7 | 14.9 | 2.7 | 14.4 | 3.6 | 0.189 |
| Gender | Women | 100 | 58% | 34 | 49% | 10* | 100% | 0.009 |
|  | Men | 72 | 42% | 35 | 51% | 0 | 0% |  |
| Race | White | 92 | 53% | 25* | 36% | 6 | 60% | 0.022 |
|  | Black | 80 | 47% | 44 | 64% | 4 | 40% |  |
| eGFR (mL/min/1.73m<sup>2</sup>) |  | 79 | 19.5 | 71* | 22.2 | 67.2 | 15.8 | <0.001 |
| APOE-ε4 carrier | Yes | 46 | 27% | 21 | 30% | 7* | 70% | 0.021 |
|  | No | 122 | 71% | 43 | 62% | 3 | 30% |  |
|  | Missing | 4 | 2% | 5 | 7% | 0 | 0% |  |
| Neuroimaging |  |  |  |  |  |  |  |  |
| Gray Matter volume /ICV |  | 0.281 | 0.021 | 0.268** | 0.020 | 0.263* | 0.023 | <0.001 |
| Hippocampus/ICVx100 |  | 0.683 | 0.111 | 0.644* | 0.116 | 0.571* | 0.140 | 0.007 |
| Cortical Thickness (mm) |  | 2.688 | 0.109 | 2.637* | 0.136 | 2.573* | 0.150 | 0.002 |
| InWMH/ICV |  | -13.166 | 1.297 | -12.899 | 1.680 | -12.488 | 1.116 | 0.366 |
| Total WMH Volume (ml) |  | 5.321 | 6.197 | 8.725* | 9.833 | 8.409 | 8.431 | 0.017 |
| GM Free Water Fraction |  | 0.199 | 0.032 | 0.211* | 0.043 | 0.240** | 0.048 | 0.001 |
| WM Free Water Fraction |  | 0.148 | 0.021 | 0.150 | 0.028 | 0.162 | 0.036 | 0.103 |
| Microbleeds present | No | 106 | 66% | 38 | 58% | 5 | 56% | 0.681 |
|  | Yes | 54 | 34% | 27 | 42% | 4 | 44% |  |
| Microbleeds count | 0 | 106 | 66% | 40 | 60% | 5 | 56% | 0.325 |
|  | 1 | 32 | 20% | 12 | 18% | 4 | 44% |  |
|  | 2+ | 22 | 14% | 15 | 22% | 0 | 0% |  |
| Lacune present | No | 112 | 70% | 40 | 60% | 6 | 67% | 0.193 |
|  | Yes | 48 | 30% | 27 | 40% | 3 | 33% |  |
| Aβ PET | Neg | 90 | 74% | 21** | 44% | 1** | 14% | <0.001 |
|  | Pos | 32 | 26% | 27 | 56% | 6 | 86% |  |

Notes: CU = cognitively unimpaired; MCI = mild cognitive impairment; Dem = probable dementia; eGFR = estimated glomerular filtration rate; Cortical Thickness = cortical thickness in areas prone to age-related dementias; GM = gray matter; WM = white matter; ICV = Intracranial volume; WMH = white matter hyperintensities; InWMH = log-normalized WMH volume (scaled by ICV); PET = positron emission tomography. Pairwise comparison p-values of significance relative to CN: \* $p < 0.05$ and \*\* $p < 0.001$ .

### Plasma AD biomarker differences across cognitive groups

We next investigated differences in cognitive status for each of the plasma AD biomarkers (**Table 2**). Relative to CU participants, those with dementia exhibited a lower Aβ42/Aβ40 ratio, and higher GFAP, p-tau217 and p-tau231 levels. Additionally, MCI participants demonstrated higher p-tau217 levels compared to CU. We did not observe statistical differences in the levels of NfL or p-tau181 across groups.

**Table 2.** Adjusted Models for each AD Plasma Biomarker.

|  | MCI vs. CU<br>(n = 66) |  |  | Dementia vs. CU<br>(n = 10) |  |  |
| --- | --- | --- | --- | --- | --- | --- |
|  | Odds Ratio | 95% CI | P-value | Odds Ratio | 95% CI | P-value |
| Aβ42/Aβ40 | 0.524 | (0.199, 1.384) | 0.192 | 0.027 | (0.003, 0.263) | 0.002 |
| GFAP | 1.183 | (0.762, 1.839) | 0.454 | 3.842 | (1.539, 9.591) | 0.004 |
| NfL | 1.510 | (0.958, 2.379) | 0.076 | 2.523 | (0.892, 7.134) | 0.081 |
| p-tau181 | 1.160 | (0.896, 1.502) | 0.261 | 1.294 | (0.794, 2.110) | 0.301 |
| p-tau231 | 1.157 | (0.752, 1.780) | 0.507 | 2.223 | (1.103, 4.479) | 0.025 |
| p-tau217 | 1.499 | (1.036, 2.168) | 0.032 | 2.389 | (1.272, 4.487) | 0.007 |

Odds Ratio and 95% CI relative to CU (n=167). n=8 participants did not have all plasma biomarker data and are not included. We standardized distributions of each biomarker by log2 transformation. Models adjusted for Age at Exam 6, Years of Education, Race, Gender, and eGFR.

### Plasma AD biomarker associations with imaging outcomes

**Table 3** shows the associations between plasma AD biomarkers and imaging biomarkers of neurodegeneration and neuroinflammation in Model 2 (Model 1 presented in **Supplementary Table 2**). In Model 2, lower Aβ42/Aβ40 was associated with lower GMV and hippocampal volume, higher GFAP was associated with lower hippocampal volume, and higher NfL was associated with lower GMV and hippocampal volume, and with higher GM Free Water. Higher p-tau217 was associated with lower GMV in Model 2, and additionally associated with lower temporal cortical thickness in Model 1. Higher p-tau181 was associated with lower temporal cortical thickness only in Model 1. Plasma measures of p-tau231 were not associated with imaging biomarkers of neurodegeneration and neuroinflammation in this sample in either model. When excluding participants with CKD (**Supplementary Table 3**), associations of Aβ42/Aβ40 and p-tau217 with GMV, and of GFAP and NfL with hippocampal volume, were no longer significant; however, associations of p-tau181 and p-tau217 with hippocampal volume became significant.

**Table 3.** AD Plasma Biomarkers and Imaging Biomarkers of Neurodegeneration and Neuroinflammation.

|  | GMV/ICV |  |  | Hippocampus/ICV |  |  | Cortical Thickness |  |  | GM Free Water |  |  |
| --- | --- | --- | --- | --- | --- | --- | --- | --- | --- | --- | --- | --- |
|  | (n = 232) |  |  | (n = 232) |  |  | (n = 232) |  |  | (n = 219) |  |  |
|  | B | SE | P-value | B | SE | P-value | B | SE | P-value | B | SE | P-value |
| Aβ42/Aβ40 | 0.009 | 0.004 | 0.039 | 0.072 | 0.020 | <0.001 | -0.009 | 0.024 | 0.720 | -0.010 | 0.007 | 0.132 |
| GFAP | -0.002 | 0.002 | 0.345 | -0.019 | 0.009 | 0.045 | 0.011 | 0.011 | 0.290 | 0.003 | 0.003 | 0.303 |
| NfL | -0.007 | 0.002 | <0.001 | -0.021 | 0.010 | 0.032 | -0.005 | 0.011 | 0.642 | 0.009 | 0.003 | 0.006 |
| p-tau181 | -0.001 | 0.001 | 0.340 | -0.009 | 0.006 | 0.094 | -0.009 | 0.006 | 0.154 | 0.003 | 0.002 | 0.166 |
| p-tau231 | -0.002 | 0.002 | 0.181 | -0.012 | 0.009 | 0.158 | -0.003 | 0.010 | 0.744 | 0.005 | 0.003 | 0.074 |
| p-tau217 | -0.004 | 0.001 | 0.018 | -0.014 | 0.007 | 0.063 | -0.017 | 0.009 | 0.052 | 0.002 | 0.002 | 0.422 |

Model 2 adjusted for Age at Exam 6, Years of Education, Race, Gender, Smoking Status, eGFR, *APOE*- ε4, and BMI. n=8 participants did not have all plasma biomarker data and are not included. We standardized distributions of each biomarker by log2 transformation.

**Table 4** shows the associations between plasma AD biomarkers and imaging biomarkers of cerebral small vessel disease in Model 2 (Model 1 presented in **Supplementary Table 4**).

**Table 4.** AD Plasma Biomarkers and Imaging Biomarkers of cSVD.

|  | InWMH/ICV<br>(n = 231) |  |  | WM Free Water<br>(n = 224) |  |  | Cerebral microbleeds<br>(n = 231) |  |  | Lacunar infarction<br>(n = 233) |  |  |
| --- | --- | --- | --- | --- | --- | --- | --- | --- | --- | --- | --- | --- |
|  | B | SE | P-value | B | SE | P-value | Odds Ratio | 95% CI | P-value | Odds Ratio | 95% CI | P-value |
| Aβ42/Aβ40 | -0.382 | 0.265 | 0.151 | -0.009 | 0.005 | 0.064 | 0.386 | (0.153, 0.976) | 0.044 | 0.789 | (0.308, 2.021) | 0.621 |
| GFAP | 0.272 | 0.120 | 0.024 | 0.002 | 0.002 | 0.284 | 0.764 | (0.504, 1.157) | 0.204 | 1.067 | (0.698, 1.632) | 0.765 |
| NfL | 0.267 | 0.126 | 0.034 | 0.005 | 0.002 | 0.022 | 1.246 | (0.815, 1.906) | 0.309 | 1.284 | (0.826, 1.998) | 0.267 |
| p-tau181 | 0.088 | 0.072 | 0.221 | 0.003 | 0.001 | 0.020 | 0.886 | (0.684, 1.148) | 0.359 | 0.927 | (0.721, 1.19) | 0.551 |
| p-tau231 | 0.143 | 0.109 | 0.190 | 0.005 | 0.002 | 0.021 | 1.474 | (0.996, 2.18) | 0.052 | 1.085 | (0.736, 1.6) | 0.680 |
| p-tau217 | 0.149 | 0.096 | 0.120 | 0.005 | 0.002 | 0.007 | 1.376 | (0.979, 1.933) | 0.066 | 1.124 | (0.797, 1.585) | 0.506 |

Model 2 adjusted for Age at Exam 6, Years of Education, Race, Gender, Smoking Status, eGFR, *APOE*- ε4, and BMI. n=8 participants did not have all plasma biomarker data and are not included. We standardized distributions of each biomarker by log2 transformation. Odds Ratio and 95% CI relative to CMB = 0 (n = 151), Lacune = 0 (n = 161)

Higher NfL was associated with higher WMH volume and WM Free Water in Model 2, and additionally associated with greater prevalence of lacunar infarcts in Model 1. All three plasma p-tau measurements (p-tau181, p-tau217, p-tau231) were positively associated with WM Free Water but not with other cerebral small vessel disease biomarkers in both models. Lower Aβ42/Aβ40 ratio was significantly associated with greater prevalence of microbleeds in Model 2. Higher GFAP was associated with higher WMH volume in Model 2. When excluding participants with CKD (**Supplementary Table 5**), associations of GFAP with WMH volume, NfL with WMH volume and WM Free Water, and p-tau231 with WM Free Water, were no longer significant.

**Table 5** shows associations between plasma AD biomarkers and imaging biomarkers of Aβ deposition (CL) and Aβ positivity in the subset with PET, in Model 2. As anticipated, lower plasma Aβ42/Aβ40 ratio was associated with higher Aβ deposition and higher odds of Aβ positivity, and higher p-tau217 and p-tau231 were also associated with higher Aβ deposition and odds of Aβ positivity, such that a doubling in the level of p-tau217 and p-tau231 were associated with a 2-3-fold increase in the odds of Aβ PET positivity. Additionally, in Model 1 (presented in **Supplementary Table 6**), higher GFAP was associated with higher Aβ deposition. These associations remained unchanged when excluding n=44 participants with CKD (**Supplementary Table 7**).

**Table 5.** AD Plasma Biomarkers and Amyloid PET

| | Centiloids | | | Aβ+ ($\geq 12.2$ CL) | | |
| --- | --- | --- | --- | --- | --- | --- |
|  | (n = 177) |  |  | (n = 177) |  |  |
|  | B | SE | P-value | Odds Ratio | 95% CI | P-value |
| Aβ42/Aβ40 | -34.221 | 6.986 | < 0.001 | 0.031 | (0.007, 0.142) | < 0.001 |
| GFAP | 5.831 | 3.409 | 0.089 | 1.173 | (0.695, 1.978) | 0.550 |
| NfL | 5.050 | 3.795 | 0.185 | 1.192 | (0.66, 2.152) | 0.560 |
| p-tau181 | 1.136 | 1.988 | 0.569 | 1.12 | (0.833, 1.507) | 0.452 |
| p-tau231 | 8.507 | 2.892 | 0.004 | 2.214 | (1.25, 3.922) | 0.006 |
| p-tau217 | 11.869 | 2.419 | < 0.001 | 2.635 | (1.542, 4.503) | < 0.001 |

Model 2 adjusted for Age at Exam 6, Years of Education, Race, Gender, Smoking Status, eGFR, *APOE*- ε4, and BMI. n=8 participants did not have all plasma biomarker data and are not included. We standardized distributions of each biomarker by log2 transformation. Odds Ratio and 95% CI relative to Aβ - (<12.2 CL; n = 113)

## Discussion

While the prevailing consensus is that these plasma biomarkers represent measures of AD pathophysiology or related neurodegeneration, we show that GFAP, NfL, p-tau181, p-tau217, and p-tau231 *also* are associated with vascular comorbidities and imaging biomarkers of cSVD. Interestingly, WM Free Water and to a lesser extent WMH were associated with multiple plasma biomarker levels. Specifically, higher plasma NfL levels were most consistently associated with measures of cSVD including higher burden of WMH and WM Free Water and a greater odds of lacunar infarction. Along with NfL, GFAP was also significantly associated with WMH in this sample. We observed consistent positive associations of all p-tau isoforms (e.g., 181, 217, 231) with higher WM Free Water. Generally, lacunar infarcts were not associated with plasma biomarker levels with one exception for presence of lacunar infarcts being associated with higher NfL levels. In contrast, plasma amyloid biomarker abnormalities, characterized by lower Aβ42/Aβ40 ratio, follow a “classic AD pathology” through associations with *APOE*-ε4 carriage, lower GM and hippocampal volumes, greater amyloid deposition, and dementia, without vascular biomarker involvement aside from a notable association with greater presence of cerebral microbleeds.

This work provides insights into the associations between plasma AD biomarkers and participant demographics and health characteristics. Importantly, plasma biomarker levels were not significantly different between Black and White participants in this study. Plasma biomarkers showed more broad involvement with vascular comorbidities and biomarkers. GFAP levels appeared to be influenced by age, gender, and comorbidities such as obesity and kidney function, and were associated with hippocampal volume, WMH, and cognitive status. NfL levels appeared to be influenced by age, kidney function and smoking and broadly associated with gray matter structure (e.g., total GM, hippocampal volume, and GM Free Water). Generally, p-tau isoforms showed weak and non-significant associations with kidney function and no associations with participant characteristics including *APOE*-ε4 carriage. P-tau217 and p-tau231 showed clear associations with AD-related biomarkers, including greater amyloid deposition, lower GMV (for p-tau217), and lower cortical thickness in temporal lobe meta-ROI, areas prone to age-related dementia. Of note, none of the p-tau levels were associated with age in this sample, although the age range is somewhat limited. Interestingly, smoking status was associated with most plasma AD biomarker levels including Aβ42/Aβ40 ratio, NFL, and all p-tau isoforms, but not with GFAP. These associations will need to be investigated further in larger samples.

In addition, when excluding participants with eGFR≤60, some associations with imaging biomarkers of neurodegeneration, neuroinflammation, and cSVD were tempered, suggesting that the relationships we observed may have been accentuated in participants with CKD. However, it is notable that associations with amyloid PET (Centiloids or positivity) were unchanged when excluding participants with CKD, strengthening our confidence in the associations of plasma biomarkers with amyloid PET.

The observed associations between plasma AD biomarkers and participant demographics and comorbidities are consistent with prior studies^12^, suggesting that age, kidney function, and smoking status impacted biomarker levels. Recently, similar associations were reported for plasma NfL with WMH and temporal lobe atrophy, but not microbleeds or lacunar infarcts^52^. In contrast, Qu et al. reported associations between higher NfL and the presence of microbleeds^53^ while also reporting associations with lacunar infarcts, WMH, and total cSVD burden. These associations between cSVD and tau pathology are also supported by neuropathology studies. In 982 deceased individuals with *ex vivo* MRI, WMH was associated with greater tau-tangle pathology but not amyloid, while evidence of arteriolosclerosis in the posterior watershed areas was associated with higher tau pathological changes^54^.

### Limitations

This work from the MESA study represents data from a single site, with a sample size limited to those participants with neuroimaging, cognitive adjudication, and plasma AD biomarker data.

The cohort assessed was comprised of participants who self-reported as Black and White and thus may not be generalizable to Hispanic and Chinese American participants recruited at other MESA sites. In addition, this work does not represent all forms of cSVD. While arteriosclerosis is a common form of cSVD observed on neuropathology, imaging biomarkers of arteriosclerosis are in development and not widely available or validated. The current study also lacks direct measures of tau aggregation with PET, which was not collected in this cohort.

### Future directions

These results call for further studies on the pathophysiological mechanisms underlying differences in the AD blood biomarkers. Plasma and MRI biomarker measurements are being collected across all sites and racial and ethnic groups within MESA. In future work, expanding on the current study, we plan to replicate these results in a larger and more diverse cohort of self-reported White, Black, Hispanic, and Chinese American participants of MESA.

## Conclusions

This work provides important observations about the potential relationships between cSVD and plasma biomarkers which are being developed for AD biomarker panels. It clearly shows that plasma biomarkers of NfL and p-tau are associated with imaging biomarkers of AD and cSVD. Plasma p-tau and Aβ42/40 are associated with AD pathological biomarkers across age groups, ethnicities and comorbidity burden. These results are confirmatory for NfL and may be novel for p-tau isoforms. They suggest that these plasma biomarkers may not only associate with AD pathology but also with evidence of cSVD.

## Data Availability

All data produced in the present study are available upon reasonable request to the authors

## Author Contributions

SNL and TMH conceived, designed, and executed the project. Data analysis and interpretation were carried out by SNL, CLS, JT, FGO, PRK, MH, SRH, NJA, MMM, RK, MDR, CTW, KDH, SC, TCR, KMH, SRR, BCS, HZ, KB, TKK, and TMH. SNL and TMH compiled the manuscript, which was approved by all authors.

## Acknowledgement

This research was supported by contracts HHSN268201500003I, N01-HC-95159, N01-HC-95160, N01-HC-95161, N01-HC-95162, N01-HC-95163, N01-HC-95164, N01-HC-95165, N01-HC-95166, N01-HC-95167, N01-HC-95168, and N01-HC-95169 from the National Heart, Lung, and Blood Institute; by grants UL1-TR-000040, UL1-TR-001079, and UL1-TR-001420 from the National Center for Advancing Translational Sciences (NCATS); and by grants P30AG049638, P30AG072947, T32AG033534 and R01AG054069 from the National Institute on Aging (NIA). The authors thank the other investigators, the staff, and the participants of the MESA study for their valuable contributions. A full list of participating MESA investigators and institutions can be found at http://www.mesa-nhlbi.org. The funding sources had no involvement in study design; in the collection, analysis and interpretation of data; in the writing of the report; or in the decision to submit the article for publication. The authors declare that no competing interests or conflicts of interest exist.

## References

1. Ashton NJ. Differential roles of Aβ42/40, p-tau231 and p-tau217 for Alzheimer’s trial selection and disease monitoring. Nature Medicine. 2022;28.

2. Karikari TK, Pascoal TA, Ashton NJ, et al. Blood phosphorylated tau 181 as a biomarker for Alzheimer’s disease: a diagnostic performance and prediction modelling study using data from four prospective cohorts. The Lancet Neurology. 2020;19(5):422–433. doi:10.1016/S1474-4422(20)30071-5

3. Palmqvist S, Janelidze S, Quiroz YT, et al. Discriminative Accuracy of Plasma Phospho-tau217 for Alzheimer Disease vs Other Neurodegenerative Disorders. JAMA. 2020;324(8):772–781. doi:10.1001/jama.2020.12134

4. Ashton NJ, Pascoal TA, Karikari TK, et al. Plasma p-tau231: a new biomarker for incipient Alzheimer’s disease pathology. Acta Neuropathol. 2021;141(5):709–724. doi:10.1007/s00401-021-02275-6

5. Ashton NJ, Janelidze S, Al Khleifat A, et al. A multicentre validation study of the diagnostic value of plasma neurofilament light. Nat Commun. 2021;12(1):3400. doi:10.1038/s41467-021-23620-z

6. Benedet AL, Milà-Alomà M, Vrillon A, et al. Differences Between Plasma and Cerebrospinal Fluid Glial Fibrillary Acidic Protein Levels Across the Alzheimer Disease Continuum. JAMA Neurology. 2021;78(12):1471–1483. doi:10.1001/jamaneurol.2021.3671

7. Teunissen CE, Verberk IMW, Thijssen EH, et al. Blood-based biomarkers for Alzheimer’s disease: towards clinical implementation. The Lancet Neurology. 2021;21(1):66–77. doi:10.1016/S1474-4422(21)00361-6

8. Karikari TK, Ashton NJ, Brinkmalm G, et al. Blood phospho-tau in Alzheimer’s disease: analysis, interpretation, and clinical utility. Nature Reviews Neurology. Published online 2022. doi:10.1038/s41582-022-00665-2

9. Nakamura A, Kaneko N, Villemagne VL, et al. High performance plasma amyloid-β biomarkers for Alzheimer’s disease. Nature. 2018;554(7691):249-254. doi:10.1038/nature25456

10. Moscoso A, Grothe MJ, Ashton NJ, et al. Longitudinal Associations of Blood Phosphorylated Tau181 and Neurofilament Light Chain With Neurodegeneration in Alzheimer Disease. JAMA Neurol. 2021;78(4):396. doi:10.1001/jamaneurol.2020.4986

11. Lim YY, Maruff P, Kaneko N, et al. Plasma Amyloid-β Biomarker Associated with Cognitive Decline in Preclinical Alzheimer’s Disease. JAD. 2020;77(3):1057–1065. doi:10.3233/JAD-200475

12. Ramanan VK, Graff-Radford J, Syrjanen J, et al. Association of Plasma Biomarkers of Alzheimer Disease With Cognition and Medical Comorbidities in a Biracial Cohort.

13. O’Bryant SE, Petersen M, Hall J, Johnson LA. Medical comorbidities and ethnicity impact plasma Alzheimer’s disease biomarkers: Important considerations for clinical trials and practice.

14. Ashton NJ, Moseby-Knappe M, Benedet AL, et al. Alzheimer Disease Blood Biomarkers in Patients With Out-of-Hospital Cardiac Arrest.

15. Rubenstein R, Chang B, Yue JK, et al. Comparing Plasma Phospho Tau, Total Tau, and Phospho Tau–Total Tau Ratio as Acute and Chronic Traumatic Brain Injury Biomarkers. Published online 2017.

16. Graff-Radford J, Mielke MM, Hofrenning EI, et al. Association of plasma biomarkers of amyloid and neurodegeneration with cerebrovascular disease and Alzheimer’s disease. Neurobiol Aging. 2022;119:1–7. doi:10.1016/j.neurobiolaging.2022.07.006

17. Romero JR, Demissie S, Beiser A, et al. Relation of plasma beta-amyloid, clusterin, and tau with cerebral microbleeds: Framingham Heart Study. Ann Clin Transl Neurol. 2020;7(7):1083–1091. doi:10.1002/acn3.51066

18. McCarter SJ, Lesnick TG, Lowe VJ, et al. Association Between Plasma Biomarkers of Amyloid, Tau, and Neurodegeneration with Cerebral Microbleeds. J Alzheimers Dis. 2022;87(4):1537–1547. doi:10.3233/JAD-220158

19. Shir D, Graff-Radford J, Hofrenning EI, et al. Association of plasma glial fibrillary acidic protein (GFAP) with neuroimaging of Alzheimer’s disease and vascular pathology. Alzheimers Dement (Amst). 2022;14(1):e12291. doi:10.1002/dad2.12291

20. Mielke MM, Frank RD, Dage JL, et al. Comparison of Plasma Phosphorylated Tau Species With Amyloid and Tau Positron Emission Tomography, Neurodegeneration, Vascular Pathology, and Cognitive Outcomes. JAMA Neurol. 2021;78(9):1108. doi:10.1001/jamaneurol.2021.2293

21. Frantellizzi V, Pani A, Ricci M, Locuratolo N, Fattapposta F, De Vincentis G. Neuroimaging in Vascular Cognitive Impairment and Dementia: A Systematic Review. Pastori D, ed. JAD. 2020;73(4):1279–1294. doi:10.3233/JAD-191046

22. Schindler SE, Karikari TK, Ashton NJ, Henson RL, Yarasheski KE. Effect of race on prediction of brain amyloidosis by plasma Aβ42/Aβ40, phosphorylated tau, and neurofilament light. Neurology. 2022;in press. doi:10.1212/WNL.0000000000200358

23. Howell JC, Watts KD, Parker MW, et al. Race modifies the relationship between cognition and Alzheimer’s disease cerebrospinal fluid biomarkers. Alzheimer’s Research & Therapy. 2017;9(1):88. doi:10.1186/s13195-017-0315-1

24. Garrett SL, McDaniel D, Obideen M, et al. Racial Disparity in Cerebrospinal Fluid Amyloid and Tau Biomarkers and Associated Cutoffs for Mild Cognitive Impairment. JAMA Netw Open. 2019;2(12). doi:10.1001/jamanetworkopen.2019.17363

25. Morris JC, Schindler SE, McCue LM, et al. Assessment of Racial Disparities in Biomarkers for Alzheimer Disease. JAMA Neurol. 2019;76(3):264–273. doi:10.1001/jamaneurol.2018.4249

26. Kumar VV, Huang H, Zhao L, et al. Baseline Results: The Association Between Cardiovascular Risk and Preclinical Alzheimer’s Disease Pathology (ASCEND) Study. Journal of Alzheimer’s Disease. 2020;75(1):109–117. doi:10.3233/JAD-191103

27. Deters KD, Napolioni V, Sperling RA, et al. Amyloid PET Imaging in Self-Identified Non-Hispanic Black Participants of the Anti-Amyloid in Asymptomatic Alzheimer’s Disease (A4) Study. Neurology. 2021;96(11):e1491–e1500. doi:10.1212/WNL.0000000000011599

28. Meeker KL, Wisch JK, Hudson D, et al. Socioeconomic Status Mediates Racial Differences Seen Using the AT(N) Framework. Annals of Neurology. 2021;89(2):254–265. 10.1002/ana.25948

29. Moonen JEF, Nasrallah IM, Detre JA, et al. Race, sex, and mid-life changes in brain health: Cardia MRI substudy. Alzheimer’s & Dementia. 2022;n/a(n/a). doi:10.1002/alz.12560

30. O’Bryant SE, Zhang F, Petersen M, et al. Neurodegeneration from the AT(N) framework is different among Mexican Americans compared to non-Hispanic Whites: A Health & Aging Brain among Latino Elders (HABLE) Study. *Alzheimer’s & Dementia: Diagnosis*, Assessment & Disease Monitoring. 2022;14(1):e12267. doi:10.1002/dad2.12267

31. Xiong C, Luo J, Schindler SE, et al. Racial differences in longitudinal Alzheimer’s disease biomarkers among cognitively normal adults. Alzheimer’s & Dementia. 2022;n/a(n/a). doi:10.1002/alz.12608

32. Kitaguchi N, Tatebe H, Sakai K, et al. Influx of Tau and Amyloid-β Proteins into the Blood During Hemodialysis as a Therapeutic Extracorporeal Blood Amyloid-β Removal System for Alzheimer’s Disease. Journal of Alzheimer’s Disease. 2019;69(3):687–707. doi:10.3233/JAD-190087

33. Nho K, Kueider-Paisley A, Ahmad S, et al. Association of Altered Liver Enzymes With Alzheimer Disease Diagnosis, Cognition, Neuroimaging Measures, and Cerebrospinal Fluid Biomarkers. JAMA Netw Open. 2019;2(7):e197978. doi:10.1001/jamanetworkopen.2019.7978

34. Mielke MM. Performance of plasma phosphorylated tau 181 and 217 in the community. Nature Medicine. 2022;28.

35. Syrjanen JA, Vemuri P, Knopman DS, et al. Associations of amyloid and neurodegeneration plasma biomarkers with comorbidities.

36. Olson JL, Bild DE, Kronmal RA, Burke GL. Legacy of MESA. Glob Heart. 2016;11(3):269–274. doi:10.1016/j.gheart.2016.08.004

37. Bild DE, Bluemke DA, Burke GL, et al. Multi-Ethnic Study of Atherosclerosis: objectives and design. Am J Epidemiol. 2002;156(9):871–881.

38. Levey AS, Coresh J, Greene T, Marsh J, Stevens LA, Kusek JW. Expressing the Modification of Diet in Renal Disease Study Equation for Estimating Glomerular Filtration Rate with Standardized Serum Creatinine Values. Clinical Chemistry. 2007;(4).

39. Ding J, Lohman K, Molina A, et al. The association between aging-related monocyte transcriptional networks and comorbidity burden: the Multi-Ethnic Study of Atherosclerosis (MESA). GeroScience. 2023;45(1):197–207. doi:10.1007/s11357-022-00608-1

40. Liang S, Steffen LM, Steffen BT, et al. APOE genotype modifies the association between plasma omega-3 fatty acids and plasma lipids in the Multi-Ethnic Study of Atherosclerosis (MESA). Atherosclerosis. 2013;228(1):181–187. doi:10.1016/j.atherosclerosis.2013.02.004

41. Weintraub S, Besser L, Dodge HH, et al. Version 3 of the Alzheimer Disease Centers’ Neuropsychological Test Battery in the Uniform Data Set (UDS). Alzheimer Disease and Associated Disorders. 2018;32(1):10–17. doi:10.1097/WAD.0000000000000223

42. Sachs BC, Steenland K, Zhao L, et al. Expanded Demographic Norms for Version 3 of the Alzheimer Disease Centers’ Neuropsychological Test Battery in the Uniform Data Set. Alzheimer Disease & Associated Disorders. 2020;34(3):191–197. doi:10.1097/WAD.0000000000000388

43. Gonzalez-Ortiz F, Ferreira PCL, González-Escalante A, et al. A novel ultrasensitive assay for plasma p-tau217: Performance in individuals with subjective cognitive decline and early Alzheimer’s disease. Alzheimer’s & Dementia. 2024;20(2):1239–1249. doi:10.1002/alz.13525

44. Lockhart SN, Schaich CL, Craft S, et al. Associations among vascular risk factors, neuroimaging biomarkers, and cognition: Preliminary analyses from the Multi-Ethnic Study of Atherosclerosis (MESA). Published online 2023.

45. Schwarz CG, Gunter JL, Wiste HJ, et al. A large-scale comparison of cortical thickness and volume methods for measuring Alzheimer’s disease severity. Neuroimage Clin. 2016;11:802–812. doi:10.1016/j.nicl.2016.05.017

46. Coffin C, Suerken CK, Bateman JR, et al. Vascular and microstructural markers of cognitive pathology. Alz & Dem Diag Ass & Dis Mo. 2022;14(1). doi:10.1002/dad2.12332

47. Wardlaw JM, Smith EE, Biessels GJ, et al. Neuroimaging standards for research into small vessel disease and its contribution to ageing and neurodegeneration. Lancet Neurol. 2013;12(8):822–838. doi:10.1016/S1474-4422(13)70124-8

48. Klunk WE, Engler H, Nordberg A, et al. Imaging brain amyloid in Alzheimer’s disease with Pittsburgh Compound-B. Ann Neurol. 2004;55(3):306–319. doi:10.1002/ana.20009

49. Klunk WE, Koeppe RA, Price JC, et al. The centiloid project: Standardizing quantitative amyloid plaque estimation by PET. Alz Dement. Published online October 28, 2014. doi:10.1016/j.jalz.2014.07.003

50. La Joie R, Ayakta N, Seeley WW, et al. Multi-site study of the relationships between ante mortem [11C]PIB-PET Centiloid values and post mortem measures of Alzheimer’s disease neuropathology. Alz Dement. Published online July 8, 2018:1–41.

51. Benjamini, Yoav Y, Hochberg Y. Controlling the False Discovery Rate: A Practical and Powerful Approach to Multiple Testing. Journal of the Royal Statistical Society: Series B (Methodological). 1995;57(1). 10.1111/j.2517-6161.1995.tb02031.x

52. Chong JR, Hilal S, Ashton NJ, et al. Brain atrophy and white matter hyperintensities are independently associated with plasma neurofilament light chain in an Asian cohort of cognitively impaired patients with concomitant cerebral small vessel disease.

53. Qu Y, Tan CC, Shen XN, et al. Association of Plasma Neurofilament Light With Small Vessel Disease Burden in Nondemented Elderly.

54. Kapasi A. Association of small vessel disease with tau pathology. Acta Neuropathologica. Published online 2022.

