## Supplementary Tables for "Plasma and neuroimaging biomarkers of small vessel disease and Alzheimer’s disease in a diverse cohort: MESA"

**Supplementary Table 1. Covariates by Plasma Biomarker**

| | | Aβ42/Aβ40 | | | GFAP | | | NfL | | | p-tau181 | | | p-tau217 | | | p-tau231 | | |
|---|---|---|---|---|---|---|---|---|---|---|---|---|---|---|---|---|---|---|---|
| | | **B** | **SE** | **P-value** | **B** | **SE** | **P-value** | **B** | **SE** | **P-value** | **B** | **SE** | **P-value** | **B** | **SE** | **P-value** | **B** | **SE** | **P-value** |
| Median Age Split | < 70 y | | Ref | | | Ref | | | Ref | | | Ref | | | Ref | | | Ref | |
| | > 70 y | -0.52 | 0.20 | 0.0096 | 35.99 | 11.19 | 0.0015 | 12.71 | 4.73 | 0.0077 | -21.97 | 23.46 | 0.3499 | -0.57 | 0.95 | 0.5506 | -4.51 | 8.12 | 0.5788 |
| Education (y) | | 0.04 | 0.03 | 0.2124 | 1.29 | 1.96 | 0.5095 | 0.47 | 0.83 | 0.5695 | -2.47 | 4.12 | 0.5492 | -0.11 | 0.17 | 0.5185 | -0.85 | 1.42 | 0.5531 |
| eGFR | | 0.00 | 0.00 | 0.8576 | -0.72 | 0.27 | 0.0091 | -0.57 | 0.11 | <.0001 | -0.64 | 0.57 | 0.2680 | -0.04 | 0.02 | 0.0742 | -0.32 | 0.20 | 0.1136 |
| Gender | Women | 0.22 | 0.19 | 0.2542 | 38.08 | 10.85 | 0.0005 | -2.86 | 4.58 | 0.5336 | 15.00 | 22.82 | 0.5115 | 0.53 | 0.93 | 0.5647 | 4.38 | 7.90 | 0.5799 |
| | Men | | Ref | | | Ref | | | Ref | | | Ref | | | Ref | | | Ref | |
| Race | White | | Ref | | | Ref | | | Ref | | | Ref | | | Ref | | | Ref | |
| | Black | -0.13 | 0.20 | 0.5179 | 9.59 | 11.04 | 0.3858 | 1.74 | 4.66 | 0.7093 | 22.80 | 23.24 | 0.3276 | 1.07 | 0.94 | 0.2556 | 9.41 | 8.01 | 0.2414 |
| BMI | | 0.03 | 0.02 | 0.0855 | -3.52 | 0.99 | 0.0005 | -0.57 | 0.42 | 0.1787 | -0.91 | 2.15 | 0.6703 | -0.10 | 0.09 | 0.2704 | -0.41 | 0.73 | 0.5715 |
| Current Smoker | No | | Ref | | | Ref | | | Ref | | | Ref | | | Ref | | | Ref | |
| | Yes | -0.90 | 0.38 | 0.0169 | 0.57 | 21.18 | 0.9786 | 25.27 | 8.95 | 0.0051 | 178.24 | 45.97 | 0.0001 | 6.77 | 1.82 | 0.0002 | 55.52 | 15.50 | 0.0004 |
| *APOE*-ε4 carrier | Missing | -0.61 | 0.54 | 0.2538 | -34.48 | 30.21 | 0.2549 | -9.46 | 12.76 | 0.4590 | 2.26 | 60.33 | 0.9701 | 0.38 | 2.45 | 0.8774 | 1.24 | 20.91 | 0.9527 |
| | Yes | -0.46 | 0.21 | 0.0271 | 4.97 | 11.62 | 0.6694 | -5.93 | 4.91 | 0.2280 | -15.05 | 24.49 | 0.5394 | -0.27 | 0.99 | 0.7825 | -4.03 | 8.45 | 0.6344 |
| | No | | Ref | | | Ref | | | Ref | | | Ref | | | Ref | | | Ref | |

**Supplementary Table 2. AD Plasma Biomarkers and Imaging Biomarkers of Neurodegeneration and Neuroinflammation**

| | GMV/ICV (n = 232) | | | Hippocampus/ICV (n = 232) | | | Cortical Thickness (n = 232) | | | GM Free Water (n =219) | | |
|---|---|---|---|---|---|---|---|---|---|---|---|---|
| | B | SE | P-value | B | SE | P-value | B | SE | P-value | B | SE | P-value |
| Aβ42/Aβ40 | 0.0079 | 0.0040 | 0.050 | 0.0674 | 0.0196 | < 0.001 | -0.0046 | 0.0233 | 0.843 | -0.0102 | 0.0066 | 0.125 |
| GFAP | -0.0020 | 0.0018 | 0.271 | -0.0191 | 0.0091 | 0.036 | 0.0031 | 0.0106 | 0.770 | 0.0037 | 0.0030 | 0.220 |
| NfL | -0.0062 | 0.0018 | < 0.001 | -0.0170 | 0.0090 | 0.060 | -0.0170 | 0.0104 | 0.105 | 0.0069 | 0.0029 | 0.019 |
| p-tau181 | -0.0011 | 0.0011 | 0.305 | -0.0084 | 0.0053 | 0.116 | -0.0132 | 0.0061 | 0.032 | 0.0031 | 0.0017 | 0.078 |
| p-tau231 | -0.0025 | 0.0016 | 0.118 | -0.0123 | 0.0081 | 0.130 | -0.0119 | 0.0094 | 0.206 | 0.0051 | 0.0027 | 0.055 |
| p-tau217 | -0.0033 | 0.0014 | 0.019 | -0.0123 | 0.0069 | 0.078 | -0.0213 | 0.0080 | 0.008 | 0.0022 | 0.0023 | 0.337 |

Model 1 adjusted for Age at Exam 6, Years of Education, Race, and Gender. n=8 participants did not have all plasma biomarker data and are not included. We standardized distributions of each biomarker by log2 transformation.

**Supplementary Table 3. AD Plasma Biomarkers and Imaging Biomarkers of Neurodegeneration and Neuroinflammation, excluding CKD**

| | GMV/ICV (n = 194) | | | Hippocampus/ICV (n = 194) | | | Cortical Thickness (n = 194) | | | GM Free Water (n = 184) | | |
|---|---|---|---|---|---|---|---|---|---|---|---|---|
| | B | SE | P-value | B | SE | P-value | B | SE | P-value | B | SE | P-value |
| Aβ42/Aβ40 | 0.0071 | 0.0048 | 0.138 | 0.0646 | 0.0225 | 0.005 | -0.0210 | 0.0256 | 0.413 | -0.0072 | 0.0075 | 0.337 |
| GFAP | -0.0015 | 0.0021 | 0.477 | -0.0195 | 0.0100 | 0.054 | 0.0106 | 0.0113 | 0.348 | 0.0028 | 0.0033 | 0.408 |
| NfL | -0.0074 | 0.0022 | < 0.001 | -0.0190 | 0.0106 | 0.075 | -0.0049 | 0.0119 | 0.681 | 0.0068 | 0.0034 | 0.048 |
| p-tau181 | -0.0018 | 0.0012 | 0.160 | -0.0134 | 0.0059 | 0.025 | -0.0131 | 0.0066 | 0.050 | 0.0026 | 0.0019 | 0.188 |
| p-tau231 | -0.0022 | 0.0019 | 0.242 | -0.0119 | 0.0091 | 0.191 | 0.0006 | 0.0102 | 0.952 | 0.0026 | 0.0030 | 0.391 |
| p-tau217 | -0.0029 | 0.0017 | 0.077 | -0.0161 | 0.0079 | 0.043 | -0.0102 | 0.0089 | 0.255 | 0.0006 | 0.0026 | 0.809 |

Excluding n=44 ppts with CKD defined by eGFR<=60. Model 2 adjusted for Age at Exam 6, Years of Education, Race, Gender, Smoking Status, eGFR, *APOE*-ε4, and BMI. n=8 participants did not have all plasma biomarker data and are not included. We standardized distributions of each biomarker by log2 transformation.

**Supplementary Table 4. AD Plasma Biomarkers and Imaging Biomarkers of cSVD**

| | lnWMH/ICV (n = 231) | | | WM Free Water (n = 224) | | | Cerebral microbleeds (n = 231) | | | Lacunar infarction (n = 233) | | |
|---|---|---|---|---|---|---|---|---|---|---|---|---|
| | B | SE | P-value | B | SE | P-value | Odds Ratio | 95% CI | P-value | Odds Ratio | 95% CI | P-value |
| Aβ42/Aβ40 | -0.3888 | 0.2576 | 0.133 | -0.0069 | 0.0049 | 0.161 | 0.459 | (0.193, 1.091) | 0.078 | 0.683 | (0.284, 1.642) | 0.394 |
| GFAP | 0.2286 | 0.1158 | 0.05 | 0.0013 | 0.0022 | 0.563 | 0.744 | (0.501, 1.106) | 0.144 | 1.142 | (0.768, 1.699) | 0.511 |
| NfL | 0.2494 | 0.1155 | 0.032 | 0.0044 | 0.0022 | 0.041 | 1.177 | (0.804, 1.722) | 0.403 | 1.522 | (1.026, 2.259) | 0.037 |
| p-tau181 | 0.1164 | 0.0671 | 0.084 | 0.0025 | 0.0012 | 0.043 | 0.871 | (0.678, 1.119) | 0.281 | 1.046 | (0.83, 1.318) | 0.702 |
| p-tau231 | 0.1571 | 0.1045 | 0.134 | 0.0042 | 0.0019 | 0.030 | 1.349 | (0.946, 1.922) | 0.098 | 1.283 | (0.891, 1.846) | 0.181 |
| p-tau217 | 0.1515 | 0.0897 | 0.093 | 0.0039 | 0.0017 | 0.021 | 1.264 | (0.931, 1.715) | 0.133 | 1.267 | (0.927, 1.732) | 0.138 |

Model 1 adjusted for Age at Exam 6, Years of Education, Race, and Gender. n=8 participants did not have all plasma biomarker data and are not included. We standardized distributions of each biomarker by log2 transformation. Odds Ratio and 95% CI relative to CMB = 0 (n = 151), Lacune = 0 (n = 161)

**Supplementary Table 5. AD Plasma Biomarkers and Imaging Biomarkers of cSVD, excluding CKD**

| | lnWMH/ICV (n = 193) | | | WM Free Water (n = 189) | | | Cerebral microbleeds (n = 193) | | | Lacunar infarction (n = 195) | | |
|---|---|---|---|---|---|---|---|---|---|---|---|---|
| | B | SE | P-value | B | SE | P-value | Odds Ratio | 95% CI | P-value | Odds Ratio | 95% CI | P-value |
| Aβ42/Aβ40 | -0.4333 | 0.3019 | 0.153 | -0.0080 | 0.0055 | 0.152 | 0.331 | (0.116, 0.944) | 0.039 | 0.685 | (0.236, 1.988) | 0.487 |
| GFAP | 0.2513 | 0.1327 | 0.060 | 0.0027 | 0.0025 | 0.274 | 0.816 | (0.518, 1.284) | 0.379 | 0.995 | (0.622, 1.591) | 0.983 |
| NfL | 0.1760 | 0.1408 | 0.213 | 0.0034 | 0.0025 | 0.190 | 1.212 | (0.760, 1.934) | 0.420 | 1.23 | (0.760, 1.991) | 0.398 |
| p-tau181 | 0.0857 | 0.0792 | 0.281 | 0.0028 | 0.0014 | 0.048 | 0.883 | (0.660, 1.181) | 0.402 | 0.862 | (0.650, 1.144) | 0.303 |
| p-tau231 | 0.1388 | 0.1201 | 0.249 | 0.0034 | 0.0022 | 0.123 | 1.316 | (0.878, 1.973) | 0.183 | 1.051 | (0.694, 1.590) | 0.815 |
| p-tau217 | 0.1544 | 0.1053 | 0.144 | 0.0051 | 0.0019 | 0.007 | 1.218 | (0.849, 1.748) | 0.284 | 1.006 | (0.698, 1.449) | 0.975 |

Excluding n=44 ppts with CKD defined by eGFR<=60. Model 2 adjusted for Age at Exam 6, Years of Education, Race, Gender, Smoking Status, eGFR, *APOE*-ε4, and BMI. n=8 participants did not have all plasma biomarker data and are not included. We standardized distributions of each biomarker by log2 transformation. Odds Ratio and 95% CI relative to CMB = 0 (n = 124), Lacune = 0 (n = 137).

**Supplementary Table 6. AD Plasma Biomarkers and Amyloid PET**

| | Centiloids (n = 177) | | | Aβ+ (≥12.2 CL) (n = 177) | | |
|---|---|---|---|---|---|---|
| | B | SE | P-value | Odds Ratio | 95% CI | P-value |
| Aβ42/Aβ40 | -37.5428 | 6.7992 | < 0.001 | 0.032 | (0.008, 0.128) | < 0.001 |
| GFAP | 7.2330 | 3.4482 | 0.037 | 1.318 | (0.836, 2.077) | 0.234 |
| NfL | 6.1558 | 3.7135 | 0.099 | 1.305 | (0.795, 2.143) | 0.293 |
| p-tau181 | 1.3778 | 2.0047 | 0.493 | 1.135 | (0.876, 1.471) | 0.336 |
| p-tau231 | 9.4910 | 2.9346 | 0.001 | 2.378 | (1.367, 4.137) | 0.002 |
| p-tau217 | 12.9945 | 2.3584 | < 0.001 | 3.023 | (1.822, 5.016) | < 0.001 |

Model 1 adjusted for Age at Exam 6, Years of Education, Race, and Gender. n=8 participants did not have all plasma biomarker data and are not included. We standardized distributions of each biomarker by log2 transformation. Odds Ratio and 95% CI relative to Aβ- (<12.2 CL; n = 113).

**Supplementary Table 7. AD Plasma Biomarkers and Amyloid PET, excluding CKD**

| | Centiloids (n = 147) | | | Aβ+ (≥12.2 CL) (n = 147) | | |
|---|---|---|---|---|---|---|
| | B | SE | P-value | Odds Ratio | 95% CI | P-value |
| Aβ42/Aβ40 | -37.946 | 7.644 | < 0.001 | 0.018 | (0.003, 0.107) | < 0.001 |
| GFAP | 4.599 | 3.723 | 0.219 | 1.217 | (0.682, 2.17) | 0.507 |
| NfL | 7.231 | 4.165 | 0.085 | 1.559 | (0.81, 3.001) | 0.184 |
| p-tau181 | 1.462 | 2.070 | 0.481 | 1.125 | (0.826, 1.534) | 0.455 |
| p-tau231 | 6.920 | 3.072 | 0.026 | 2.125 | (1.158, 3.899) | 0.015 |
| p-tau217 | 11.097 | 2.499 | < 0.001 | 2.38 | (1.371, 4.133) | 0.002 |

Excluding n=44 ppts with CKD defined by eGFR<=60. Model 2 adjusted for Age at Exam 6, Years of Education, Race, Gender, Smoking Status, eGFR, APOE-ε4, and BMI. n=8 participants did not have all plasma biomarker data and are not included. We standardized distributions of each biomarker by log2 transformation. Odds Ratio and 95% CI relative to Aβ- (<12.2 CL; n = 96).
